# Primed for Exploitation: How Early Violence, Institutional Betrayal, and Structural Vulnerability Shape Pathways into Pornography

**DOI:** 10.64898/2026.05.07.26352588

**Authors:** Meghan Donevan, Inga Dennhag, Carl Göran Svedin, Jennifer Martin, Linda S. Jonsson

**Affiliations:** Department of Clinical Sciences, Child and Adolescent Psychiatry, Umeå University, Umeå, Sweden; Department of Social Sciences, Marie Cederschiöld University, Stockholm, Sweden; Faculty of Community Services, Toronto Metropolitan University, Toronto, Canada; Centre for Combating Child Sex Trafficking and Online Child Sexual Exploitation, Toronto Metropolitan University, Toronto, Canada; Talita, Stockholm, Sweden

**Keywords:** Pornography, child sexual abuse, institutional betrayal, socio-ecological framework, sexual exploitation, mixed methods

## Abstract

Women filmed for pornography report extensive abuse and serious health consequences, yet pathways into pornography remain under-examined. Using an embedded qualitative mixed-methods approach, we explored factors shaping these pathways in Sweden. Twenty-five adults (23 women) who had been filmed for pornography completed questionnaires and participated in teller-focused interviews. Informed by a socio-ecological framework, our reflexive thematic analysis generated the global theme *Primed for exploitation*, comprising three themes: *Imprints of early violence*, *No one has my back: Relational and institutional betrayals*, and *Compounding structural vulnerabilities*. Our findings reveal how childhood abuse and violence, relational and institutional betrayals, material precarity, and a pornified cultural landscape converge to shape pathways into pornography. To prevent and disrupt these pathways, early identification of sexual abuse, timely access to trauma-informed care that avoids individualizing and pathologizing the consequences of violence, and practical support that addresses material precarity are critical. From a socio-ecological perspective, framing entry into pornography as a simple matter of “choice” is fundamentally flawed: it individualizes deeply social processes and obscures the profound impact of cumulative violence, repeated relational and institutional betrayals, and intersecting structural constraints.

## Introduction

The production of pornography is increasingly recognized as a site of significant harm, where vulnerabilities are leveraged and abuse is recorded and sold for profit. Survivor accounts, criminal investigations, civil litigation, and government inquiries have documented pervasive violence and exploitation across the industry, with young women and girls disproportionately affected.^1–3^ Mounting evidence also indicates that those filmed for pornography experience serious adverse mental health outcomes, including posttraumatic stress and suicidality.^3, 4–8^ Taken together, these realities position being filmed for pornography as an important women’s health concern with substantial consequences for mental, physical, sexual, and relational health. This underscores the need to understand the pathways and conditions that lead women and others into pornography in the first place.

Previous research consistently links being filmed for pornography with intersecting and cumulative vulnerabilities such as poverty, histories of abuse, mental health challenges, race and gender discrimination, and social marginalization.^6, 7, 9–12^ Adverse childhood experiences are particularly prevalent: across diverse studies and settings, individuals filmed for pornography report high rates of childhood abuse and placement in institutional or foster care.^4, 7, 9, 13^ For example, a Swedish study among 120 individuals (88% women) filmed for pornography found that 96% were subjected to some form of child abuse (88% sexual abuse, 90% psychological abuse, and 79% physical abuse), and over one-third had been placed in institutional or foster care.^4^ Global research on youth affected by sexual exploitation further demonstrates how multiple co-occurring disadvantages in childhood predict later exploitation.^9, 13, 14, 15^

While earlier research has established common risk factors and vulnerabilities among individuals filmed for pornography, quantitative studies alone cannot fully capture how these factors interact to shape an individual’s pathway into pornography. Qualitative research, especially lived-experience studies, is better suited to illuminating processes, meanings, and contextual dynamics. Evidence from related fields illustrates this. A qualitative study on street prostitution, for instance, demonstrates how the combined impact of childhood sexual exploitation and the failure of social and welfare systems to identify, protect and support vulnerable youth can consolidate pathways into street prostitution as adults.^16^ Similar dynamics were observed in qualitative research on intimate partner violence, where researchers found that earlier victimization, the resulting isolation and loneliness, and dismissive or harmful responses when seeking help were critical factors in young women’s trajectories into abusive relationships.^17^

Pathways into pornography may echo such patterns, but they are also likely shaped by dynamics specific to today’s digital, cultural, and economic landscape. One example is the normalization of platforms like OnlyFans, a subscription-based platform where pornographic content is bought and sold. Promoted as a source of empowerment and financial freedom,^18^ the mainstreaming of such platforms can obscure exploitative practices and normalize sexual objectification in ways that may be particularly influential in shaping young women’s pathways. Additionally, the newness of these platforms means that earlier research does not adequately capture how risk factors and vulnerabilities unfold within today’s emerging forms of pornography distribution.

This gap underscores the urgency of examining how pathways into pornography emerge, evolve, and might be disrupted. Using a qualitatively driven mixed-methods approach, this study examines factors shaping trajectories into pornography through questionnaires and in-depth interviews with adults who have been filmed for pornography. By foregrounding participants’ lived experiences while integrating quantitative indicators, this study offers new insight into the mechanisms through which exploitation pathways are formed. These insights have direct implications for prevention, early identification, and the development of trauma-informed support interventions.

### Conceptualizing Pornography

In this study, we focus on *commercial* pornography, which we define as sexual acts that are livestreamed, recorded, or otherwise documented for real-time or delayed viewing in exchange for compensation. This compensation (monetary or material) may be received by the individuals depicted or may accrue partially or entirely to third parties. In some instances, images are captured by the individual depicted and sold directly to buyers (i.e., “self-produced”). More commonly, third parties—including traffickers, so-called “pornography producers,” prostitution buyers, and platform operators—are involved and exert varying degrees of control and derive profit from the content. Our definition intentionally excludes image-based sexual abuse that does not involve compensation, as these crimes (e.g., AI-generated pornography or a partner non-consensually sharing a sexual image) may represent isolated incidents of abuse rather than ongoing commercial exploitation.

This conceptualization foregrounds the role of third parties (profiteers), including digital platforms, while situating pornography within established frameworks on sexual exploitation and gender-based violence. Our operationalization recognizes pornography’s continuity with other forms of commercial sexual exploitation, encompassing technology-facilitated prostitution and trafficking for sexual exploitation.^19–21^

### Theoretical Starting Point

Given the high prevalence of childhood abuse among those filmed for pornography, trauma-informed theoretical frameworks provide a useful starting point for understanding why early traumatic experiences, particularly childhood sexual abuse, can produce enduring patterns of vulnerability to exploitation.^22–25^ Finkelhor and Browne’s^26^ traumagenic dynamics model demonstrates how traumatic sexualization, betrayal, powerlessness, and stigmatization associated with child sexual abuse shape children’s self-concept and relational expectations, instilling the belief that they are fundamentally damaged or unworthy, and affecting their worldviews—for example, the belief that the world is unsafe.

Herman^24, 27^ builds on this framework by emphasizing how child sexual abuse fractures one’s identity. She says, “Such early violations of trust and safe attachment damage the formation of a coherent sense of self, embodied in space, continuous in time, and deserving of love.”^27 p 58^ As these children reach adulthood, “these lasting injuries compromise their ability to form relationships of trust, intimacy, and mutuality,”^27 p 59^ potentially increasing susceptibility to exploitation.

Several scholars caution against pathologizing victims and instead frame trauma symptoms and coping behaviours as intelligible responses to profoundly harmful experiences.^29–33^ It is also crucial to consider how social, institutional, and professional responses can either reinforce or alleviate the effects of violence. Such responses communicate whether the violence is acknowledged and taken seriously.^30–32^ Positive reactions validate the victim’s experience and support healing, whereas negative social responses such as disbelief, minimization, or blame can deepen the sense of betrayal and intensify suffering. Indeed, one study found that negative social reactions and lack of support can be stronger predictors of trauma severity than the original event.^33^

In other words, trauma is not confined to the abuse itself but is shaped by the wider relational, community, and structural contexts in which it occurs. This aligns with research showing that vulnerability to sexual exploitation emerges through the interaction of individual and contextual factors.^13^ To capture these multi-level influences, we draw on a social-ecological framework that situates individual experiences within broader social contexts across the individual, relationship, community, and societal levels.^35^ In the context of contemporary pornography, however, this framework must also account for the digital environment. Building on Martin and colleagues, we treat the online environment as a cross-cutting cybersystem that interacts with and influences the other ecological systems.^36, 37^ This adapted social-ecological framework guides our analysis of how individual vulnerabilities and broader social-structural conditions interact, offline and online, to shape pathways into pornography.

## Methods

The data for this study were generated as part of a larger research project commissioned by a Swedish Governmental inquiry on the Protection, Support and Care of Individuals Subjected to Abuse in the Production or Distribution of Pornography.^3^ We adopted a qualitatively driven, embedded mixed-methods design to enable a holistic and in-depth exploration of pathways into pornography.^38–40^ While the analysis prioritized qualitative inquiry, the embedded quantitative component helped contextualize participant characteristics and experiences, thereby complementing and enriching the in-depth interview data. Interviews were conducted using the teller-focused interview method^41^ and analysis was guided by reflexive thematic analysis.^42, 43^ Ethical approval was granted by the Swedish Ethical Review Authority (reference number 2022-06718-01).

### Recruitment

Participants were recruited through frontline service providers and non-governmental organizations that likely encounter the target population. Service providers distributed a research flyer describing the study and how to contact the research team. Flyers were distributed in physical service settings (e.g., offices and clinics) as well as via organizational social media accounts. While recruitment primarily targeted the greater Stockholm area, dissemination through social media facilitated broader geographic reach across Sweden.

To participate in the original study, individuals were required to be at least 18 years old and have been filmed for pornography. In total, forty individuals responded to the recruitment flyer and expressed interest in participating in the study. Those who met the inclusion criteria and wished to proceed selected the time and location for the interview. A total of 28 interviews were conducted between March 2023 and October 2023.

For the present study, we focused specifically on participants with experiences of *commercial* pornography (as defined above). Twenty-five of the 28 participants met this criterion and provided complete questionnaire and interview data.

Participants reported experiences of both self-produced and third-party-produced pornography, often across multiple contexts and over time, including instances where filming occurred within the context of conventional (in-person) prostitution. Recorded sexual acts were sold or distributed through direct messaging apps (e.g., KIK, WhatsApp), subscription platforms (e.g., OnlyFans, Scandalbeauties), mainstream tube sites (e.g., Pornhub), social media (e.g., Snapchat Premium), dating apps (e.g., Grindr), pornography producers’ own sites, BDSM forums, and prostitution advertising websites.

### Data collection

Data collection proceeded in two stages. Participants first completed a structured questionnaire and then participated in an in-depth qualitative interview. Participants selected the location for their interviews. When conducted in person (e.g., café, library, university room), a quiet and secluded area was selected to ensure privacy. Interviews could also be conducted via a secure university-provided video platform.

In the first phase, participants completed a questionnaire consisting of 168 main questions with sub-questions (total 304 questions). The questionnaire covered a range of domains, including background characteristics, childhood experiences, circumstances in pornography including exposure to violence and abuse, health status, and experiences seeking and receiving support. The questionnaire incorporated the four standardized instruments (Table 1). For the present article, we report a subset of LYLES items selected a priori as most relevant to interpersonal victimization and adverse childhood circumstances related to pathways into pornography.

**Table 1:** Questionnaire instruments completed during data collection.

| **Instrument** | **Description** |
| --- | --- |
| Linköping<br>Youth Life<br>Event Scale | An inventory of adverse events experienced by the respondent or a close relative during upbringing.<sup>44</sup> It covers 41 events (13 interpersonal, 18 non-interpersonal, 10 prolonged), including sexual abuse. Responses are binary |
| (LYLES) | (Yes/No), with higher totals indicating greater exposure to adverse events, i.e., polytraumatization. |
| PTSD Checklist (PCL-5) | A 20-item scale assessing posttraumatic stress disorder (PTSD) symptoms according to diagnostic criteria.<sup>45, 46</sup> Each item is rated on a Likert scale of 0–4 (not at all to extremely), based on experiences in the past month. Total scores range from 0–80, with four subscales reflecting the symptom clusters of PTSD. |
| Symptom Checklist-25 (SCL-25) | A 25-item scale adapted from the SCL-90<sup>47</sup> assessing psychological symptoms, primarily depression and anxiety, over the past week.<sup>48, 49</sup> Items are rated 1–4 (not at all to extremely), yielding a total score of 25–100 and two subscales (anxiety, depression). |
| Dissociation Screening Questionnaire 12 (DSQ-12) | A 12-item screening tool assessing dissociative symptoms.<sup>50</sup> Items are rated on a 1–5 frequency scale (never to always), producing a total score from 12–60. The questionnaire captures both psychoform and somatoform dissociation through two subscales. |

After completing the questionnaire, the interviewer introduced the purpose and structure of the qualitative interview. The interviews drew on the teller-focused interview method developed for research on violence and abuse.^41^ While comparable to a semi-structured interview, the method gives participants considerable space to narrate their experiences and highlight what they see as most important to them, creating space for voices often historically silenced.

An interview guide ensured coverage of several key thematic areas (e.g., childhood experiences, entrance into pornography, experiences in pornography, and seeking support and help). However, the emphasis remained on participants’ life stories, narrated in their own words and at their own pace. Sample questions included: *Can you tell me about your childhood? Can you tell me about the first time you were filmed for pornography?*

Interviews took place within a 60-minute time frame, with the possibility of follow-up sessions. Participants received an honorarium of 500 SEK (approximately 50 EUR) in appreciation for their time and insights. They were also provided with the contact details for the research team and specialized community organizations and agencies if they required support after the interview.^51^

### Ethical considerations

The interviews were guided by the principles of feminist, trauma-informed, and violence-informed research.^41, 51, 52^ These principles include prioritizing the emotional well-being of participants, communicating with warmth and compassion, seeking ways to reduce the power imbalances between researcher and participant, and, when appropriate, providing information that helps participants contextualize, understand or normalize their experiences.^41, 52^

The teller-focused interview method emphasizes the creation of a *relationally safe space*, where interviewer and participant engage as partners with distinct but complementary roles.^41^ The method acknowledges inherent power imbalances but seeks to minimize them through mutual respect, shared responsibility, and support for the participant’s narration. Participants are given the choice of interview location to enhance their sense of safety and control. Consent was also approached as an “ongoing process,”^41 p 800^ requiring situationally adapted ethics,^53, 54^ with participant well-being prioritized at all stages. We also sought to balance the collection of rich and relevant data with participant’s wish to share their stories and focus on topics most important to them.^55^

In this study, the honorarium served as another mechanism for reducing power imbalances by recognizing participants’ expertise and central role in co-creating knowledge. The selected amount (500 SEK) was chosen to acknowledge participants’ time and knowledge while remaining modest enough to avoid coercion.^56^

Consistent with previous studies showing that trauma survivors generally do not experience adverse effects from discussing past trauma,^51, 52, 57^ participants in this study described the interview as an overall positive experience. For some, it was the first time they had spoken openly about their experiences related to pornography. Several participants said the interview helped them better understand themselves and their experiences, and many were motivated to participate in order to support others facing similar situations.

### Analysis

Quantitative data were analyzed in IBM SPSS Statistics (version 29) using descriptive statistics (counts and summary statistics). Quantitative indicators were used to contextualize and triangulate themes.

Interviews were audio-recorded and transcribed verbatim. All identifiable information was removed, and pseudonyms were assigned to participants. We used reflexive thematic analysis,^42, 43^ selected for its theoretically flexible yet systematic approach. Guided by a critical realist epistemological stance, we conducted an experientially oriented analysis, examining both semantic (explicit) and latent (underlying) features of the data to capture participants’ accounts alongside our own interpretations. This approach aligned well with our aims to foreground the voices and experiences of a marginalized, historically silenced group,^39^ while critically examining the social and cultural structures that shape their accounts.

After reviewing audio-recordings and transcripts for familiarization, initial codes were applied to all data segments potentially relevant to the research aims. Codes were then collated and organized into broader candidate themes that captured key patterns in the data. Themes were iteratively reviewed and refined by the first and last authors through repeated engagement with codes and data extracts. The final themes and manuscript drafts were reviewed collectively by all authors.

All but one of the interviews were conducted in Swedish. From the coding stage onward, analysis was conducted in English. Relevant Swedish quotations were translated into English by the bilingual members of the research team and checked to ensure the meaning was captured correctly.

While cognisant of how our own prior experiences and knowledge shaped the questions asked and interpretations made, we treated researcher subjectivity as a resource in line with the principles of reflexive thematic analysis.^43^ Our collective expertise in psychology, psychiatry, and social work, together with our engagement in prior theoretical and empirical research, informed the development of relevant interview questions. Our extensive experience of conducting interviews with children and adults affected by sexual exploitation also supported rapport-building and ethical sensitivity. Several participants noted that the relevance of the questions and the interviewers’ familiarity with the topic signalled expertise, facilitating candid disclosure and more detailed accounts.

## Results

We first present descriptive questionnaire findings to contextualize the sample, followed by the qualitative themes and subthemes with illustrative quotations.

### Participant Characteristics

The sample comprised 25 individuals (23 women and 2 men) aged 18 to 38 (Table 2). Most were Swedish-born, identified as bisexual or heterosexual, and had not completed post-secondary education. Financial precarity was common: nearly half reported monthly incomes of 10,000–20,000 SEK, and several were unemployed or living in safe houses or group homes.

**Table 2.**
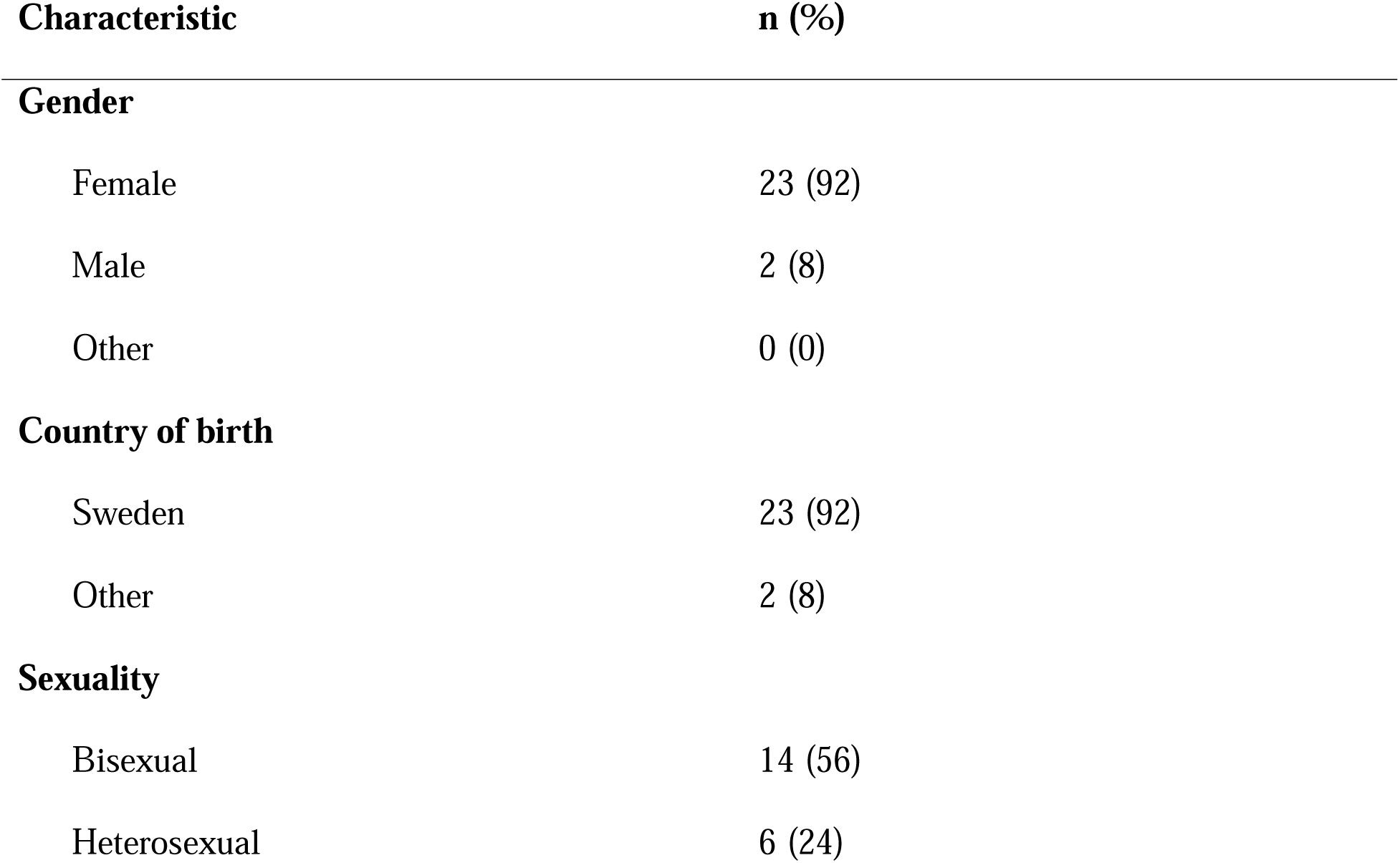
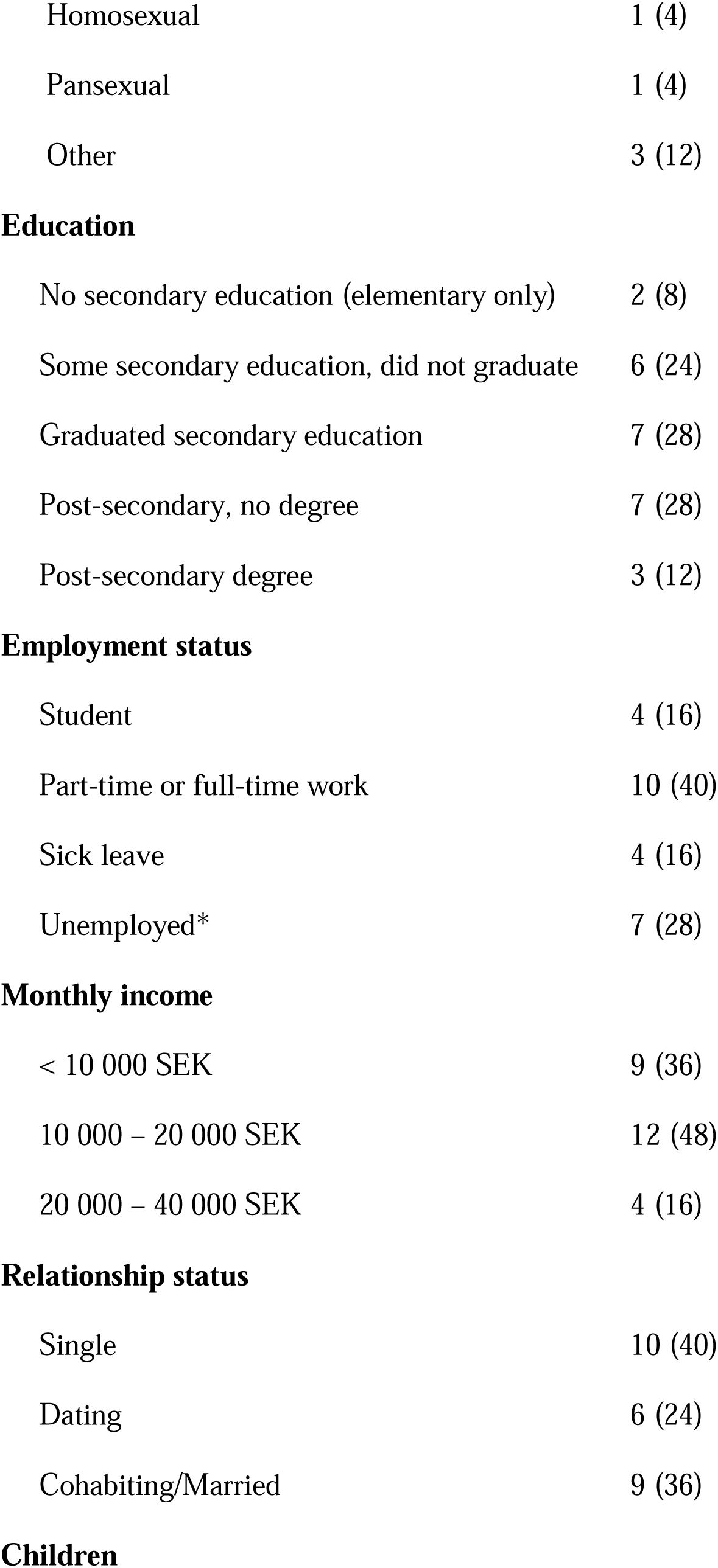
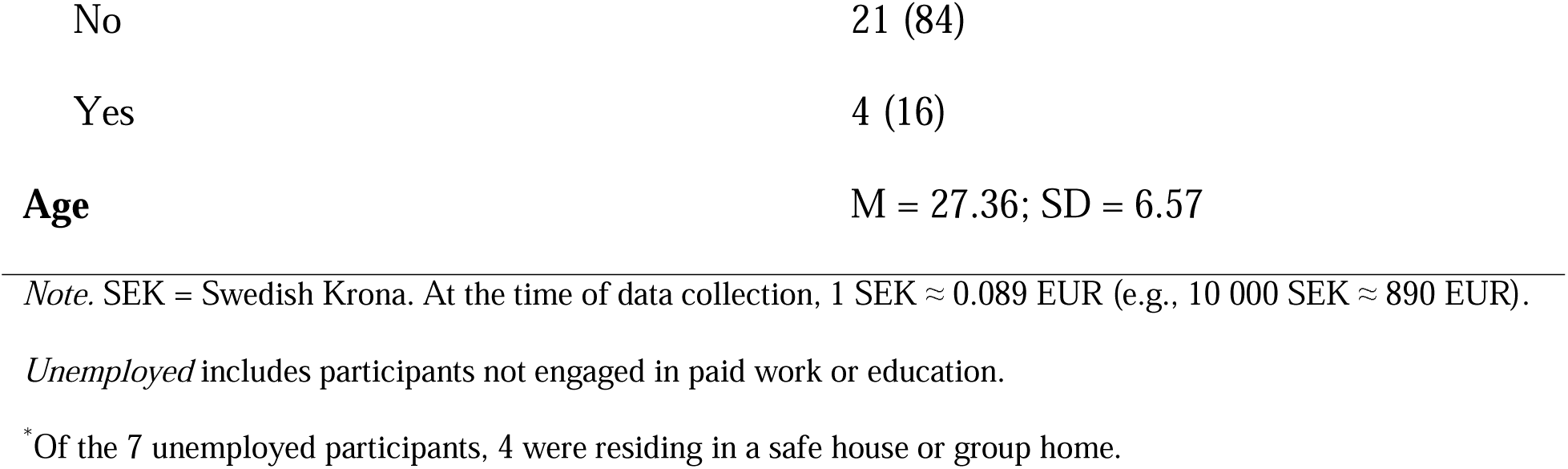
Participant Characteristics (N = 25)

| Characteristic | n (%) |
| --- | --- |
| **Gender** |  |
| Female | 23 (92) |
| Male | 2 (8) |
| Other | 0 (0) |
| **Country of birth** |  |
| Sweden | 23 (92) |
| Other | 2 (8) |
| **Sexuality** |  |
| Bisexual | 14 (56) |
| Heterosexual | 6 (24) |
| Homosexual | 1 (4) |
| Pansexual | 1 (4) |
| Other | 3 (12) |
| **Education** |  |
| No secondary education (elementary only) | 2 (8) |
| Some secondary education, did not graduate | 6 (24) |
| Graduated secondary education | 7 (28) |
| Post-secondary, no degree | 7 (28) |
| Post-secondary degree | 3 (12) |
| **Employment status** |  |
| Student | 4 (16) |
| Part-time or full-time work | 10 (40) |
| Sick leave | 4 (16) |
| Unemployed* | 7 (28) |
| **Monthly income** |  |
| < 10 000 SEK | 9 (36) |
| 10 000 – 20 000 SEK | 12 (48) |
| 20 000 – 40 000 SEK | 4 (16) |
| **Relationship status** |  |
| Single | 10 (40) |
| Dating | 6 (24) |
| Cohabiting/Married | 9 (36) |
| **Children** |  |
| No | 21 (84) |
| Yes | 4 (16) |
| **Age** | M = 27.36; SD = 6.57 |

*Note.* SEK = Swedish Krona. At the time of data collection, 1 SEK ≈ 0.089 EUR (e.g., 10 000 SEK ≈ 890 EUR).
*Unemployed* includes participants not engaged in paid work or education.
\*Of the 7 unemployed participants, 4 were residing in a safe house or group home.

### Potentially Traumatic Childhood Experiences

Table 3 shows extensive and overlapping childhood victimization and adversity, often alongside caregiver dysfunction. Sexual victimization was nearly universal, and entry into pornography typically occurred during adolescence: the mean age at first filming was 15.5 years (range 6–25), and 68% (17/25) reported that initial exploitation occurred during childhood.

**Table 3.** Potentially Traumatic Childhood Experiences (N = 25)

| **Type** | **n (%)** |
| --- | --- |
| **Victimization (by adults/peers)** |  |
| Sexual abuse by adult in family (incest) | 10 (40) |
| Sexual abuse by someone else | 24 (96) |
| Physical violence by adult in family | 15 (60) |
| Physical violence by someone else | 19 (76) |
| Emotional/psychological abuse | 23 (92) |
| Bullying by peers | 16 (64) |
| Exposure to domestic/partner violence in the home | 14 (56) |
| Threatened with harm to self or loved one | 18 (72) |
| **Household dysfunction and other adversities** |  |
| Caregiver mental health problems | 19 (76) |
| Caregiver alcohol/drug problems | 14 (56) |
| Caregiver long-term illness/disability | 13 (52) |
| Separation from caregivers (foster/institutional care) | 8 (32) |
| Participant long-term illness or disability | 14 (56) |

*Note: Table includes selected LYLES items judged most relevant to interpersonal victimization and adverse childhood circumstances in relation to the study aim. Multiple responses permitted.*

### Current psychiatric symptom burden

Scores on the three health questionnaires indicated a high current symptom burden in the sample (Table 1). Participants had a mean PCL-5 score of 46.64 (SD = 17.90), exceeding commonly used screening thresholds of 31–33 for probable PTSD.^58^ The mean SCL-25 score was 65.20 (SD = 15.76), corresponding to a mean item score of 2.61, which is above the suggested cut-off of >1.75 for clinically significant psychological distress.^59^ This score was also higher than scores reported in an earlier Swedish study among women who had no experiences of prostitution (N= 2,554, M = 34.0) and those who did (N = 16, M = 42.4).^60^ Participants also scored highly on the DSQ-12, with a mean total of 36.00 (SD = 11.36). This is substantially above both the suggested cut-off score of 25 and the mean score of 17 reported in a Swedish study of young people aged 10–20 years (N = 451).^50^ Taken together, the questionnaire results suggest elevated levels of posttraumatic stress, general psychological distress, and dissociative symptoms at the time of the study, further contextualizing the extensive psychiatric comorbidity reported by participants.

### Psychiatric diagnoses and medications

Anxiety disorders and PTSD/complex PTSD were the most commonly reported clinician-diagnosed conditions among participants (Table 4). Psychiatric comorbidity was high: participants reported a median of four diagnoses (range 0–8), and 84% had three or more. Most participants (80%) also reported current psychotropic medication use, most commonly antidepressants and anxiolytics/sedatives.

**Table 4.**
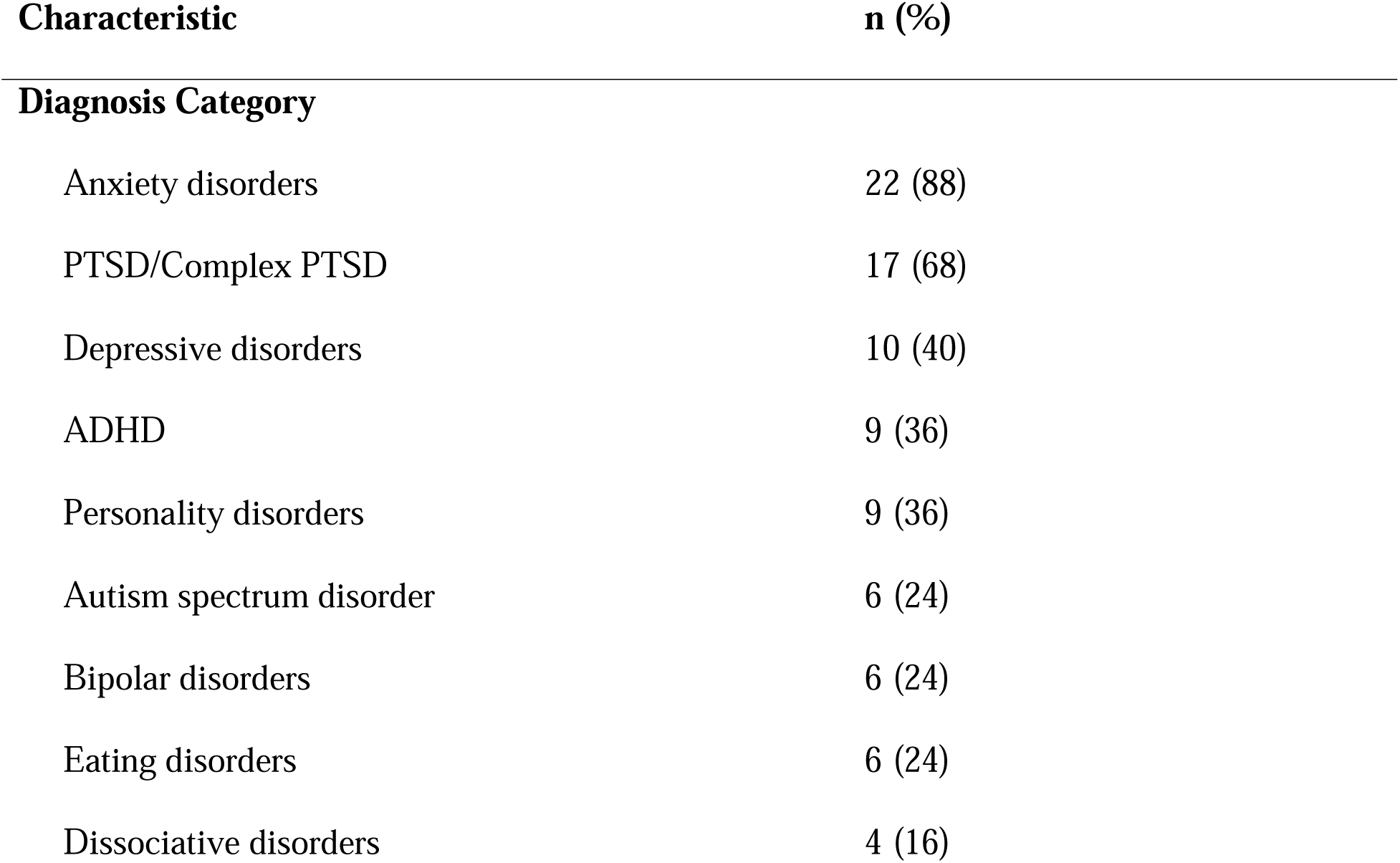
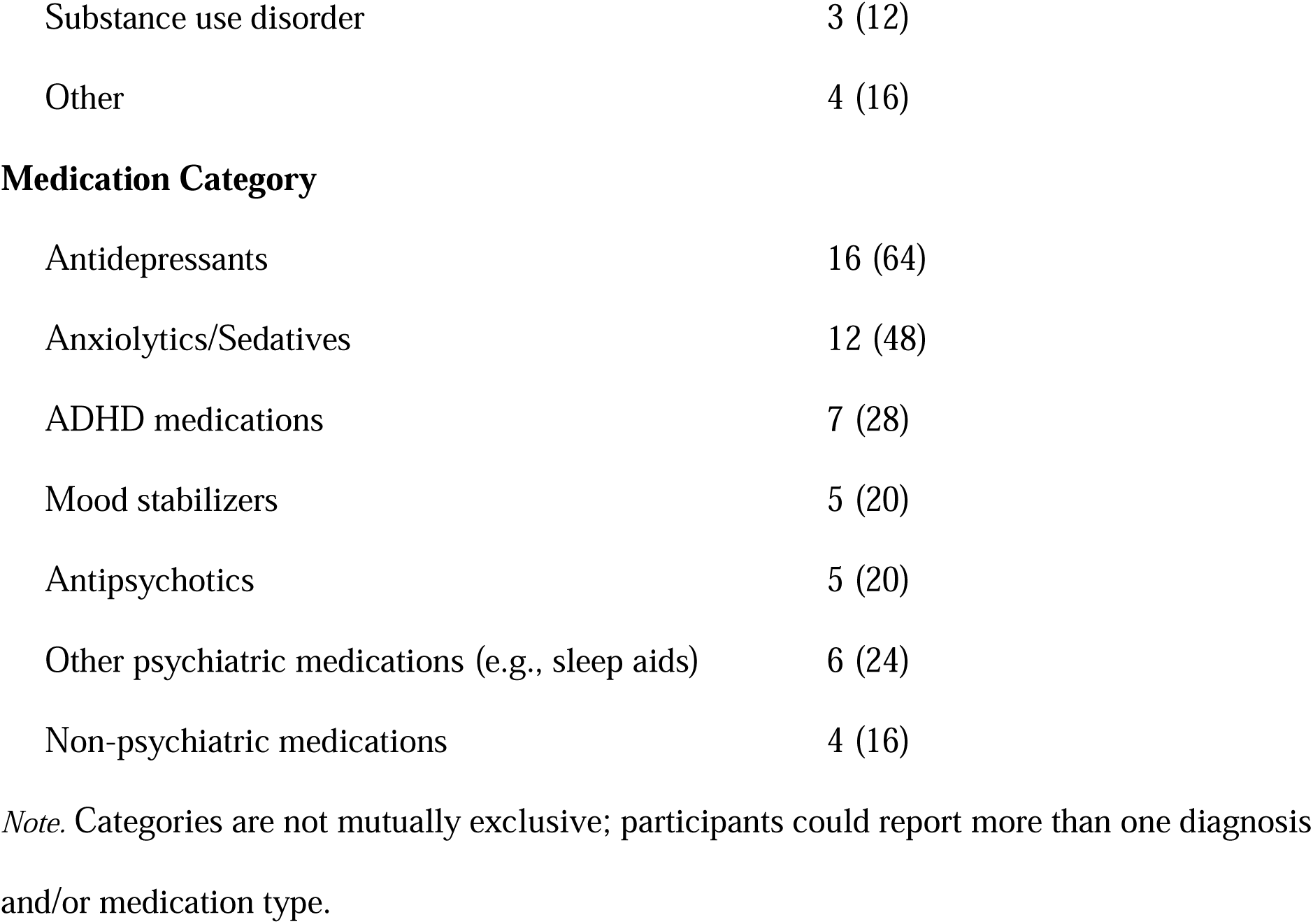
Psychiatric Diagnoses and Medication Prescriptions (N = 25)

| Characteristic | n (%) |
| --- | --- |
| **Diagnosis Category** |  |
| Anxiety disorders | 22 (88) |
| PTSD/Complex PTSD | 17 (68) |
| Depressive disorders | 10 (40) |
| ADHD | 9 (36) |
| Personality disorders | 9 (36) |
| Autism spectrum disorder | 6 (24) |
| Bipolar disorders | 6 (24) |
| Eating disorders | 6 (24) |
| Dissociative disorders | 4 (16) |
| Substance use disorder | 3 (12) |
| Other | 4 (16) |
| **Medication Category** |  |
| Antidepressants | 16 (64) |
| Anxiolytics/Sedatives | 12 (48) |
| ADHD medications | 7 (28) |
| Mood stabilizers | 5 (20) |
| Antipsychotics | 5 (20) |
| Other psychiatric medications (e.g., sleep aids) | 6 (24) |
| Non-psychiatric medications | 4 (16) |

*Note.* Categories are not mutually exclusive; participants could report more than one diagnosis and/or medication type.

### Contact with Authorities and Support Services

Finally, we asked participants which authorities and public services they had ever been in contact with or had sought support from (Table 5). We did not ask about the purpose or timing of these contacts; the aim was solely to assess the number and range of institutional contact points. Across the full sample, participants reported a median of 10 contact points (range = 2–14, M = 9.2, SD = 2.9), indicating extensive engagement with multiple institutional systems. Nearly half (48%) reported contact with ten or more services throughout the course of their lives, most commonly law enforcement, social services, and psychiatric services.

**Table 5.** Contact with Authorities and Support Services (N = 25)

| **Authority / Service** | **n (%)** |
| --- | --- |
| Law Enforcement | 22 (88) |
| Adult Psychiatry | 21 (84) |
| Social Services | 20 (80) |
| Child and Adolescent Psychiatry | 20 (80) |
| Emergency Psychiatry | 20 (80) |
| Youth Clinic | 16 (64) |
| Social Insurance Agency | 15 (60) |
| Public Employment Service | 11 (44) |
| Civil society organization | 10 (40) |
| Private therapist or psychologist | 10 (40) |
| Enforcement Authority | 9 (36) |
| Regional Specialized Support Clinic *Mikamottagningen* | 9 (36) |
| Tax Agency | 6 (24) |
| Other | 9 (36) |

*Note.* Translations of Swedish authorities and services: Adult Psychiatry (*Psykiatrin*); Emergency Psychiatry (*Akutpsykiatrin*); Child and Adolescent Psychiatry (*Barn- och ungdomspsykiatrin*; *BUP*); Social Insurance Agency (*Försäkringskassan*); Public Employment Service (*Arbetsförmedlingen*); Enforcement Authority (*Kronofogden*); Tax Agency (*Skatteverket*); and Mikamottagningen (regional specialized support service for individuals affected by prostitution).

Taken together, the quantitative data reveal a sample characterized by extensive trauma exposure, high psychiatric comorbidity, and frequent contact with mental health and social service systems, providing an important context for the qualitative themes outlined below.

### Qualitative Findings

Our qualitative analysis generated the global theme *Primed for Exploitation*, articulated through three interrelated themes: 1) *Imprints of Early Violence*, 2) *No One Has My Back: Relational and Institutional Betrayals*, and 3) *Compounding Structural Vulnerabilities.* Several of these themes include subthemes that capture distinct, yet related aspects of the theme explored. The themes and subthemes are visualized in Figure 1, and each theme is unpacked in detail below.

**Figure 1:**
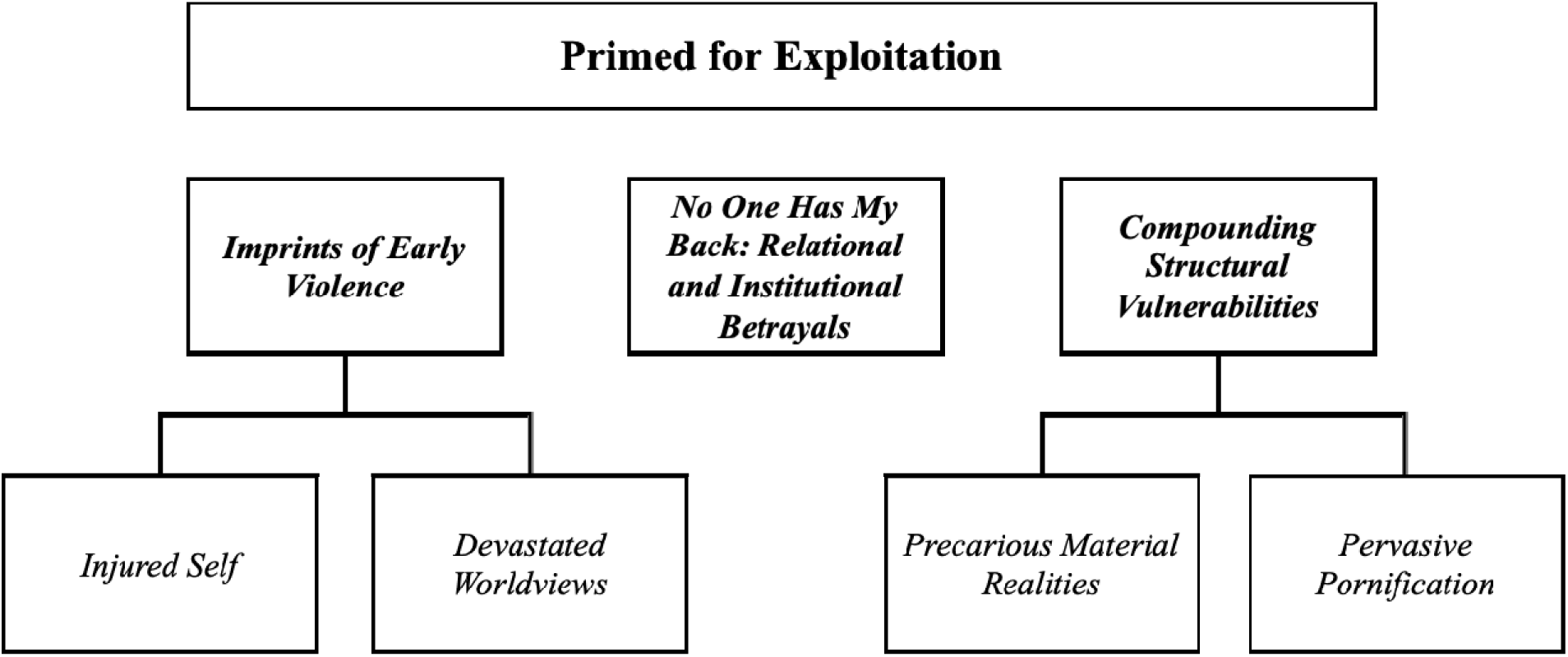
Themes and Subthemes.

### Imprints of Early Violence

This theme examines how early abuse left enduring embodied and psychological imprints that shaped participants’ pathways into pornography. When describing their childhoods, participants recounted repeated violations, chronic fear, and multiple forms of abuse. Many described growing up in a near-constant state of hypervigilance, exposed to and directly targeted by violence in ways that profoundly shaped their sense of self, their bodies, and their expectations of others.

Within this wider context of violence and polyvictimization, sexual abuse emerged as particularly salient in shaping pathways. All but one participant reported experiences of child sexual abuse, and even the participant who appeared to be the exception (answering “no” on the questionnaire) described early online sexualization by adult men during the interview. Abuse typically began in early childhood—often by biological fathers or stepfathers—or otherwise in adolescence, perpetrated by male intimate partners, peers, teachers, or strangers encountered online. For some, exploitation in pornography began concurrently with the initial abuse, where the violations were recorded and circulated online for profit. For others, pornography became an extension of earlier violence, where participants explicitly linked their trajectory into pornography with earlier sexual abuse.

Below, we unpack two closely interlinked processes stemming from earlier sexual abuse—an injured self and devastated worldviews—and how these dynamics interact to shape vulnerability to involvement in pornography.

### Subtheme 1.1: Injured Self

When participants began sharing their stories and describing their childhoods, many focused on early experiences of sexual abuse. For some, chronic victimization throughout childhood meant that there was no cohesive sense of “self” prior to the abuse—the abuse had always been a part of their lives. For others, their first violation in adolescence was described as life-shattering, fundamentally altering their self-conception. Josefin, for example, said that she felt like a completely different person after being raped:

> Everything changed when I was 15. It was the worst thing I had ever been through. Things that happened later could maybe be seen as worse, but to me, they’re not. … Everything happened so fast, but it was so painful—physically, spiritually, emotionally, and mentally. … I remember thinking, ‘Who am I now?’ I’m still the same person. Like, it was just five minutes, nothing really happened. But… it doesn’t feel like I’m the same person anymore. (Josefin)

Whether experienced as chronic or acute, sexual abuse was consistently framed as an injury to the deepest dimensions of the self. As Marcus put it: “When I explain and talk about it [the abuse], it hurts all the way down to the soul…” Such experiences represented a profound moral and identity injury, marking the onset of internalized shame and self-blame, or the intensification of pre-existing feelings of shame stemming from other forms of abuse. Perpetrator gaslighting (e.g. “It’s your fault” or “You wanted it”) further reinforced feelings of shame and self-blame.

Alongside self-blame, participants described pervasive feelings of worthlessness and self-loathing. For some, various forms of self-harm served not only to numb and cope with this emotional pain but also as self-punishment directed at bodily parts or parts of the self they perceived as having “betrayed” them. In some cases, this punishment extended to selling sexual acts (images, live video, and in-person encounters).

> … I did everything…to hurt myself as much as possible, because I hated myself … (Marcus)
>
> I was aiming for maximum destruction. I sold myself to people whom I thought could hurt me the most. (Evelina)

One participant described coming to see such exploitation as her only possible “role,” after years of abuse and being told she would never become anything else:

> Somewhere along the way I started telling myself that… this was my function. This is what I’m supposed to do in life. I’m not good at anything else. I can’t become anything. That’s what everyone around me has basically said my whole life… (Maja)

While self-betrayal and self-loathing often came to dominate participants’ accounts, this coincided with moments of resistance when participants recognized the violence for what it was and resisted the notion that they deserved it. This tension between acceptance and rejection of self-blame was accompanied by long-standing identity struggles, as participants grappled with the question, “Who am I?” when describing their lives after the abuse. This internal split also extended to their relationship with their bodies: participants described feeling as though their bodies were alien, damaged, or no longer their own. As Malin put it, “sex became part of my life as a child in a way I never wanted…that has left me feeling dirty, less worthy, like my body doesn’t belong to me.” Together, these narratives show how early, often cumulative sexual violence injured participants’ sense of self, undermined self-worth, and altered their relationship to their own bodies—imprints that also shaped how they came to view others and the world around them.

### Subtheme 1.2: Devastated Worldviews

Early experiences also fundamentally altered how participants perceived other people, relationships, and the social world at large. These experiences undermined basic assumptions about safety, trust, and care, contributing to negative beliefs about others’ intentions and the predictability of social relationships. As a result, participants described navigating the world through a lens of chronic threat anticipation, where harm was expected rather than exceptional.

> …I definitely have [negative beliefs about the world]. I’m really skeptical of other people’s intentions and convinced that everyone is dangerous and cruel. I know that’s not really true, but my experiences tell me it is. (Marcus)

Several participants described internalizing a “fawn” survival response—appeasing others to avoid further abuse—from an early age. Malin describes how these imprints affected her later in life:

> Instead of reacting with “fight,” “flight,” or “freeze,” my reaction was “please.” I figured out that if I gave people exactly what they wanted, they would usually leave me alone for the moment. And I’ve carried that pattern into adulthood, into the sex industry—whenever I’ve felt unsafe, I try to make the other person happy instead. That way they might not, or at least it might make it a bit harder for them to, hurt me. (Malin)

Sexual abuse thus constituted early conditioning in prioritizing the needs and desires of others. In this sense, sexual abuse functioned as a “training ground” for boundaries being violated. Early violations further taught participants that sex was not about their own pleasure but something transactional—where attention, affection, and love were contingent upon sexually accommodating men’s desires.

The narratives also demonstrate how early violations “revealed the truth about men”: that men were dangerous, that men would always objectify them, and that relationships with men revolved around meeting their sexual demands at the expense of one’s own needs. For example, Alice described: “I’ve become very, well, cynical toward guys. Actually, it’s been like that since I was 15”—since her first sexual violation. Likewise, Josefin reflected:

> It was basically like, “fuck all guys” … “all men are rapists.” It became this thing of, “Go ahead, prove me wrong.” (Josefin)

Drawing on earlier sexual violations, Laura concluded that men always had an ulterior motive:

> My view of men is completely destroyed…I was 100% convinced then that every man sees women as objects. Whether they’re saying it or not. that every man is, basically wants to hurt a woman in one way or another. Always gonna hurt a woman and… Yeah, that intimacy would always, that it will always feel like being used. That it would always feel like something taken away from me or having to perform something or whatever. I was convinced of those things. (Laura)

Such imprints helped frame violence as both normal and expected, making future male violations appear inevitable.

### Theme 2: No One Has My Back – Relational and Institutional Betrayals

This theme captures how repeated relational and institutional failures to recognize, respond to, and protect against abuse further shaped participants’ pathways into pornography. It reveals how trauma is not only constituted through direct victimization, but is often more profoundly shaped by the social and institutional responses surrounding—and following—that abuse.

Throughout the narratives, mothers were most often identified as the person who should have noticed, believed the child, and intervened. Instead of recognizing the violence and intervening to protect them, mothers’ own mental health struggles or victimization often hindered their ability to respond to their child’s distress. Several participants shared experiences of their mothers responding dismissively, minimizing the abuse, or even refusing to believe their disclosures.

> Telling my mother felt worse than the abuse itself—she did nothing, as if she hadn’t heard me. After that, I felt I couldn’t tell anyone. (Elin)

Some participants recalled attempts to disclose the abuse to other trusted adults. Rather than full disclosure, they described carefully testing potential recipients with small fragments of information to gauge whether the adult appeared empathetic, knowledgeable, and capable of responding appropriately. Any sign of discomfort, dismissal, or lack of engagement resulted in withdrawal and remaining silent. This process reflects a form of anticipatory self-protection, shaped by prior experiences of disbelief and betrayal.

Barriers to direct disclosure, including prior experiences of negative social responses and fear of perpetrator retaliation, led participants to adopt other, nonverbal ways of signalling distress. Nonverbal cues typically manifested as changes in behaviours, such as altered eating patterns, new risk-taking behaviours, or acting out in school. However, these cues were overlooked for the cries for help they represented, and instead mischaracterized as “problematic behaviour.” For example, Wilma shared how her attempts to communicate that home was not a safe place for her backfired. Instead of being identified and supported by the school, she was forced to spend increasing time at home:

> School was my place. My safe place. But unfortunately, I also became the class clown and was quite disruptive at school. I wanted adults to notice me; I was desperate. … I was trying to signal at school—the one place I felt safe—that I wasn’t coping at home and that something was wrong. Instead, I was shut out there as well. (Wilma)

Some participants described coping with their pain by overachieving academically and maintaining an active social life—patterns that, unfortunately, kept adults from recognizing their vulnerabilities:

> There were so many times when I reached out for help as a child, but it felt like everyone looked away. I would drop hints that I wasn’t okay, yet no one took responsibility. Maybe part of it was that I was always ‘good’ on the surface—doing well at school, having friends and activities—and that masked what was really going on. (Evelina)

These patterns extended beyond family and into institutions. Across narratives, schools were repeatedly described as settings that could potentially have identified the abuse, yet consistently failed to do so. Social services were also often present throughout participants’ childhoods; yet still the abuse typically remained invisible, unaddressed, or inadequately responded to. As a result, participants were precluded from receiving timely, trauma– and violence-informed care and protective interventions, despite considerable contact with institutions explicitly tasked with safeguarding children.

Most participants eventually had contact with Child and Adolescent Psychiatry (CAP) and were often assigned psychiatric diagnoses such as anxiety disorders, depressive disorders, PTSD, and attention-deficit/hyperactivity disorder (ADHD). While some participants described feeling relieved and validated to receive a diagnosis, others experienced the diagnostic process as misaligned with their lived realities and failing to capture the ongoing violence and unprocessed traumas underpinning their “symptoms.” Emma, for example, shared how neither her school nor CAP inquired about her home environment—where she was being systematically exploited by her father and other men. Instead, she experienced her trauma-driven responses as being medicalized and decontextualized.

> I wish someone had asked me how things were at home. … They just assumed, “She has anxiety, so she self-harms,” and never considered that something at home might be causing it. If someone had simply asked, “Wait, how are things at home?” everything might have been different. (Emma)

Professionals’ tendency to focus on behaviours and symptoms rather than underlying trauma was a recurring theme across narratives. This symptom-focused approach was experienced as invalidating and disorienting, reinforcing participants’ sense that their suffering was being misunderstood or misattributed. For example, after being sexually abused by a relative and witnessing her mother being raped by her father, Malin’s trauma responses resulted in her being characterized as the “problem child.” Malin articulated the impact of being blamed and pathologized rather than seen and understood, what she described as institutional gaslighting:

> No one ever talked about it, ever. So, when I started having emotional outbursts, I became “the problem child.” … Nobody looked deeper or asked, “Why are you acting like this?” Everyone just looked at the symptoms. And that became the truth: “You’re the problem child—you should be punished or taught to behave.” It was as if the [abuse] didn’t exist. Being gaslit from such a young age about why you feel the way you do—it does something to you. It makes you stop trusting yourself. (Malin)

Participants characterized medication as the default institutional response to this “misbehaviour”—leaving the underlying issues unaddressed.

> I’ve tried to show signs that I feel fucking awful. Can someone just do something. Instead, they increase the medication! Well, maybe that’s not actually where the problem lies. (Johanna)

In the absence of care that addressed the underlying harm, participants were left alone in their suffering and often vilified for their coping mechanisms and cries for help. Negative responses from family, school, healthcare, and social services signalled to participants that no one was there to protect them, and implicitly, that they did not deserve protection. Unable to make sense of the violations and their consequences, some participants described turning to pornography and other forms of commercial sexual acts as a means of psychological survival. Moa articulated this with striking clarity, describing how selling sexual acts functioned as a way to avoid confronting her first rape in the absence of any support:

> I know that at the beginning I did it because I didn’t get any support or help after the first rape…because I felt that if I didn’t sell myself, I would have to *feel*—I’d have to face that first rape. And I felt I couldn’t face it; I had no support, and I thought I would die. So I sold myself to keep from dying emotionally. To escape the first [assault]. (Moa)

On a deeper level, such external betrayals helped entrench participants’ pre-existing self-betrayal and devastated worldviews. When adults failed to acknowledge the abuse and help children make sense of their responses to that abuse, shame and self-hatred were magnified and negative worldviews reinforced. There was a palpable desire among participants for relational acceptance**—**to be accepted in their entirety, abuse and all, and to have their experiences recognized as violence rather than pathologized. Such recognition within trusted relationships was described as essential for making meaning of their experiences and removing self-blame and shame. Emelie, for example, described the difference her partner’s acceptance made in her healing:

> “He said, ‘It’s not you who should be ashamed.’ And those words were so incredibly important. … He wanted to be with me even though I was broken.” (Emelie)

Ida described a similar turning point in therapy, where she felt seen as a person rather than reduced to symptoms:

> …she didn’t see me as a patient… she really saw me as a human being. She saw me as a girl who had been subjected to this. She saw me as someone she personally cared about. (Ida)

In contrast, repeated betrayals from caregivers to institutional systems formed a continuum of abandonment that further eroded trust in adults and authorities, reinforcing the sense that no one had their back and contributing to a cycle of invisibility.

### Theme 3: Compounding Structural Vulnerabilities

This final theme captures how broader structural conditions shaped participants’ trajectories into pornography. When socioeconomic hardships and the normalization of pornography intersected with traumatic conditioning and systemic failures of protection, selling pornographic content online appeared to be one of the few viable, and increasingly normalized, responses to their circumstances.

### Subtheme 3.1: Precarious Material Realities

Across participants’ accounts, precarious material conditions emerged as a significant, though not exclusive, factor shaping their pathways into pornography. For some, financial hardship had been part of their traumatic childhoods. Emma described how her family’s limited means, combined with her mother’s addiction, resulted in chronic deprivation:

> My mom worked all the time when I was little and we had very little money, even though my parents actually had decent incomes. But my mom was also a heavy alcoholic and had a lot of debt, and took out tons of SMS loans. So all the money disappeared. … I went to school in dirty clothes, my siblings came in dirty clothes, we had lots of cavities, we were underweight. It was pure misery. (Emma)

Faced with ongoing abuse and persistent material hardship, Emma described turning to selling sexual acts online in an attempt to flee her trafficker, her father.

For others, the consequences of trauma undermined school performance and impeded later income-generating opportunities. Johanna, for example, linked her limited educational attainment, ill health, and precarious work to a sense that there were few viable choices available to her:

> But what else was I supposed to do? I had no qualifications, I had only completed high school. I was on sick leave, and I was bouncing between hourly temp jobs. What was I supposed to do? I felt that my life was over…that I didn’t really have much of a future. (Johanna)

Several participants emphasized not only the lack of financial resources but also the absence of reliable support, from both familial or institutional sources. Laura described how, in the absence of support from either family or the state, selling sexual acts became her only perceived means of surviving recurrent financial crises.

> …I’ve been in situations where I couldn’t pay a bill and couldn’t turn to either my family or the state. So my solution from 15 has been to sell sexual acts. Also because of my PTSD. [I sold myself] in-person at first, but then eventually online. (Laura)

Similarly, Jenny described how a convergence of mental health deterioration linked to earlier abuse, substance use, and the discontinuation of her student support left her unable to meet basic needs. In this context, selling live webcam pornography emerged as a last-resort strategy under conditions of acute material and psychological strain:

> I had no job and no income. I had been on sick leave from my studies, and when I no longer had CSN, I had no financial support and didn’t receive sickness benefits either. … My life was essentially in chaos. I was using drugs and I was in a very bad place mentally. I needed the money and couldn’t see any other way. There were probably other options, but I wasn’t in a position to get a job. … I guess it was those circumstances that made me do it [sell live webcam]. (Jenny)

Beyond acute crises, participants also described feeling compelled as adolescents to maintain the appearance of normalcy or belonging within their peer circles by attaining certain clothing, accessories or other material products. For young people already shaped by trauma and lacking validation and support, selling pornography became one of the few accessible strategies to try to keep pace with those around them.

The material reality surrounding participants was thus one in which economic struggle interacted with the lingering effects of trauma, the absence of a social safety net, and the cumulative erosion of educational, social, and occupational opportunities, creating conditions in which pornography entered their trajectories as one of the few available means of addressing their material realities.

### Subtheme 3.2: Pervasive Pornification

A final ingredient that primed participants toward pornography was the pervasive cultural normalization of pornography and the increasingly blurred boundaries between social media and pornography. *Pornification* describes the saturation of everyday digital environments with sexualized content and the framing of pornography as normal, desirable and even glamourous. These messages were especially targeted at young women and girls.

In several participant accounts, social media platforms such as YouTube and TikTok were described as a kind of “gateway” into the world of pornography. The experience of being *seen*, gaining followers, and attracting likes and comments—set against the backdrop of earlier abuse, the ensuing loneliness, and a lack of supportive relationships—represented an attempt to meet a deep need for affirmation and acceptance. As Johanna noted,

> When I posted dance videos on YouTube I got so many compliments and comments. And that was something I wanted. I craved it. I had a huge need for validation. (Johanna)

Participants also described being sexualized online from a young age by male peers or older men. That early and frequent sexualization made such messaging eventually feel expected and normalized. Laura explained:

> It’s normal for girls to receive sexual messages and pictures from a very young age, whether we want them or not. From my first device it was already happening. (Laura)

Likewise, Oskar described how repeated sexual comments from older individuals during early adolescence led to a gradual habituation to boundary violations, making such interactions feel difficult to resist or contest:

> Those comments felt kind of weird because they were from people who were much older. A lot of people called me ‘handsome’ when I was little and gave me compliments…many focused on my looks and the fact that I was young. So I experienced vulnerability, or whatever you want to call it, in that way. … you get so many comments that you have to normalize it a little. (Oskar)

Several participants described how the transition from social media visibility (likes, followers) to selling sexualized content felt like a gradual and intuitive next step. For example, after earning extra income and building up a following on YouTube, persistent requests from followers eventually convinced Oskar to shift to OnlyFans:

> When I returned to my YouTube channel people noticed that I had started training and began commenting [that I should start on OnlyFans]. At first, I laughed it off. “No, that will never happen,” I even wrote “Not gonna happen.” But eventually I gave in.

Public framing of platforms like OnlyFans as a route to fame or quick money further reinforced perceptions that selling sexual images was a seemingly viable option to not only address economic need, but even access fame and wealth:

> …it’s really tempting at first. And I don’t think anyone can honestly say it isn’t tempting. You see people getting famous from porn and others making a lot of money on OnlyFans… (Ida)

Within this pornified cultural landscape, selling sexual images emerged not as an isolated choice, but as a culturally available and increasingly normalized response to pre-existing trauma, unmet needs for validation, and acute material pressures.

## Discussion

Individuals filmed for pornography—disproportionately women—face significant risks to their health and wellbeing, warranting a closer look at how pathways into pornography take shape, evolve, and might be disrupted. Drawing on in-depth interviews and questionnaire data, this study shows how pathways into pornography are shaped through interacting vulnerabilities across ecological levels and increasingly mediated by digital environments. Early abuse left enduring imprints at the individual level; relational and institutional betrayals deepened and consolidated these vulnerabilities; and material precarity together with a pornified, digitally saturated culture made pornography appear available, normalized, and, at times, one of the few conceivable responses to participants’ circumstances. Our contribution is thus not only to identify common vulnerabilities, but to show how they accumulate and interact to produce exploitable vulnerability.

At the individual level, participants described enduring imprints of early abuse in the form of embodied shame, fractured self-concept, altered expectations of men and intimate relationships, and early conditioning into appeasement of others. These were not psychological deficits but survival strategies: responses that are rational, even necessary, in contexts of profound and repeated violence, yet which primed participants for later or continued exploitation. Sexual abuse thus functioned as a “training ground” in which boundaries were systematically violated and survival came to depend on accommodating others’ desires.

Pornography became an extension of that conditioning. When repeated violations came to signify that men would inevitably objectify, use, and invade one’s body, selling sexual acts could appear to offer some measure of control in a “man’s world,” however injurious and constrained. For some, this same dynamic became self-punitive, with violent and exploitative sexual encounters functioning as both punishment and confirmation of entrenched beliefs about worthlessness, even as the harm was perpetrated by others. In Finkelhor and Browne’s^23^ terms, these responses reflect the same underlying *dynamic of powerlessness*. Hanson’s^61^ analysis of self-blame further helps explain why later exploitation can come to feel chosen: blaming oneself may feel less threatening than confronting the reality that one was repeatedly violated, betrayed, and overpowered despite every effort to resist. At the same time, participants also described moments of recognizing the violence as wrong and resisting the notion that they deserved it. Which of these understandings took hold depended largely on the responses of others and the wider social context.

Our findings make clear that violence and its consequences are fundamentally social. How caregivers, professionals, and institutions respond to violence determines whether children’s knowledge of wrongdoing is validated and acted upon or instead left to be carried alone. Although most participants had extensive contact with multiple healthcare and social service providers across childhood and adolescence, many described never being asked direct questions about sexual victimization and circumstances at home. In the absence of such questions, children typically communicate through nonverbal, embodied means: withdrawal, self-harm, controlled eating, risk-taking, and other forms of “acting out.”^61^ Participants’ narratives suggest that these signals were frequently misread as pathology and addressed at the level of presenting symptoms (e.g., diagnosis and medication accumulation), with little attention to the traumatic and violent context generating them. When services respond primarily at the level of symptoms (e.g., escalating medication and diagnoses) without enquiry about violence, exploitation, and safety, care can become decontextualized and inadvertently reinforce shame, self-blame, and a sense of being “the problem.” As participants grappled with competing truths about themselves, responses from caregivers, professionals, and institutions emerged as pivotal in either disrupting or entrenching narratives of blame, shame, and worthlessness.^63^ Social responses are thus not peripheral to harm but constitutive of the harm itself.

Across narratives, the experience of reaching for help and finding no adult willing or able to hold their reality with them was at least as traumatizing as the original abuse.^64^ The concept of *existential loneliness*,^65^ a feeling of complete disconnection and separation from others and from the social world, helps capture the psychological and social aftermath when violence that “hurts right down to the soul” is neither named nor met with timely intervention. Victims are left alone to carry the abuse and the burden of making sense of it in isolation, often in ways that consolidate shame, self-blame, and worthlessness. Like other forms of violence, this can create a negative feedback loop where being silenced and abandoned can increase vulnerability to further exploitation, which in turn deepens isolation.^17^ Selling sexual acts, whether online or offline, can thus function as a social and psychological survival strategy: a way of managing unbearable emotional pain in a context where help is unavailable, violence is left unnamed, and victims are implicitly made to feel—both by those around them and by wider societal narratives—that they are to blame and do not deserve protection.^66^ Without timely intervention and violence– and trauma-informed support, the trajectory too often persists from childhood into adult exploitation.^16^

Finally, our findings point to societal conditions and narratives that make such survival strategies more likely to take a commercial, online form. Material precarity, limited access to stable income, and gaps in the social safety net narrow perceived alternatives, particularly when trauma-related difficulties disrupt education, employment, and housing stability. At the same time, pornification, social media, and the mainstreaming of platforms like OnlyFans render sexual self-commodification and objectification socially normalized, framed through narratives of empowerment and “easy money.” Within this landscape, pornography becomes a culturally available response to cumulative constraint and adversity.

### Clinical and practical implications

To disrupt pathways into pornography, frontline services must strengthen early identification and response systems. First, routine screening about childhood trauma and victimization (online and offline) should be standard in key points of contact: school health services, youth clinics, child and adolescent psychiatry, primary care, and social services. Professionals should not wait for disclosure—children have compelling and legitimate reasons to hesitate—but take responsibility for inquiring about violence, safety, and support needs. This is especially important for child and adolescent psychiatry, where violence-exposed children are overrepresented among service users.^67^

Second, disclosures must reliably lead to timely specialist support through clear referral pathways, active handovers, follow-up, and cross-sector coordination. Support should be holistic, addressing both the consequences of violence and practical needs such as housing and finances.

Third, clinicians and other care providers should help children, adolescents, and adults affected by violence make sense of survival strategies and trauma responses in context (e.g., self-harm, controlled eating, dissociation, substance use, risk-taking), and help children integrate their experiences.^61^ In doing so, care providers can resist individualizing harm by treating trauma-shaped survival responses as signals of violence exposure and unmet protection needs rather than as isolated pathology, acknowledging the deeply social nature of violence.^67^

Above all, our findings stress the importance of empathetic witnessing: professionals, caregivers, or other community members who can name the violence as violence, locate responsibility with the perpetrator, and help the child make meaning of both the abuse and their responses, affirming what, as the narratives show, the child often already knows: that they were not the problem, and that they did not deserve what was done to them.^61, 62^ Creating space for victim-survivors to share their experiences on their own terms, and to piece together a coherent account of their lives, helps explain why even research interviews can have therapeutic effects by offering recognition, meaning-making, and the experience of being met with dignity and care.^32^ As Herman^27^ argues, justice is not only a matter of perpetrator accountability, but also of receiving interpersonal, institutional and societal responses that recognize the harm as harm, restore dignity, and provide the protection and support that were previously denied.

### Limitations

While this study provides insight into pathways into pornography in the Swedish context, these pathways may differ in other contexts, such as low-resource settings where material insecurity and limited access to protective systems may be even more pronounced than in a social welfare state like Sweden. Still, sexual violence is widespread globally, and gaps in timely, trauma-informed support are not unique to Sweden, suggesting that several of the mechanisms identified here may be relevant beyond Sweden even if their salience and expression vary by context. The sample was also primarily recruited through service providers (directly or indirectly through social media dissemination) which may further limit transferability. Our recruitment may underrepresent individuals with relatively fewer vulnerabilities who have not sought support, but also those most vulnerable who are unable to access services or have contact with authorities (e.g., individuals controlled by traffickers). That said, the more varied Swedish sample (n=120) in a larger research study on this particular population corroborates the pronounced nature of childhood traumas among this group,^4^ while research among adolescents in Spain underscores the impact that the normalization of OnlyFans and equivalent platforms has on adolescents generally, and vulnerable ones specifically.^18^ Even if the relative weight of drivers differs across contexts, the findings point to interacting and intersecting factors at the individual, community, institutional, and societal levels, highlighting the value of a social-ecological lens for identifying mechanisms and trajectories as well as where prevention and intervention efforts can most effectively interrupt them.

## Conclusion

Our findings demonstrate how childhood abuse and violence, relational and institutional betrayals, material precarity, and a pornified cultural and digital landscape converge to shape pathways into pornography. Rather than reflecting isolated individual choices, trajectories are shaped by interacting vulnerabilities operating across the individual, relational, institutional, and societal levels, while digital environments increasingly shape the forms through which exploitation is made available, normalized, and enacted. Entry into pornography emerges as a *response* to, and embedded within, a continuum of violence and broader systems of inequality and gendered power. Without a life-course perspective on the long-term consequences of violence and betrayal, and without attention to the wider social and structural conditions shaping constraint, exploitation will continue to be mischaracterized as “individual choice” rather than socially produced vulnerability. “Choice” is a flawed unit of analysis: these are preventable, patterned pathways shaped by institutional and societal failures of safety, support, and justice. Responsibility lies not with the exploited, but with the systems that failed to protect them and the men who exploit them.

## Acknowledgments

We are deeply grateful to the individuals who participated in this study and entrusted us with their experiences. We are also grateful to Malin Jenstav, a PhD candidate at Marie Cederschiöld University, for her contribution to data collection. The broader research project from which these data were drawn was commissioned by the Swedish Government Inquiry on the Protection, Support and Care of Individuals Subjected to Abuse in the Production or Distribution of Pornography (Dir. 2022:100). We acknowledge the use of Claude, an AI-assisted tool, for language refinement during manuscript preparation. The authors carefully reviewed and edited all content for accuracy and take full responsibility for the final manuscript.

## Author Contributions

Meghan Donevan: Conceptualization, Methodology, Formal analysis, Investigation, Data curation, Writing – original draft, Writing – review & editing, Project administration, Funding acquisition.

Inga Dennhag: Writing – review & editing.

Carl Göran Svedin: Conceptualization, Methodology, Writing – review & editing, Funding acquisition.

Jennifer Martin: Writing – review & editing.

Linda S. Jonsson: Conceptualization, Methodology, Investigation, Writing – review & editing.

## Statements and declarations

### Ethical considerations

Ethical approval was granted by the Swedish Ethical Review Authority (reference number 2022-06718-01).

## Consent to participate

All participants received written and oral information about the study, including its purpose, the voluntary nature of participation, confidentiality, and their right to withdraw. Written informed consent to participate was obtained from all participants prior to data collection.

## Consent for publication

Written informed consent for publication of de-identified data, including pseudonymized interview quotations, was obtained from all participants. Identifying details have been omitted or altered to protect confidentiality.

## Data availability

The data are not publicly available due to the sensitive nature of the study and the risk of participant identification. Access to the data is restricted in accordance with ethical approval and data protection regulations.

